# Acid Suppressing Medication Use in Infants in the United States, 2002-2018

**DOI:** 10.1101/2020.08.26.20182717

**Authors:** Michael E. Johansen

## Abstract

Introduction: I attempted to determine rates of use and associations with use of acid-suppressing medications in infants under 2 years old given these are not well studied. Methods: The 2002-2018 Medical Expenditure Panel Survey (MEPS) was used for the analysis. The survey is sponsored by the Agency of Healthcare Research and Quality and is conducted on households to be nationally representative of the non-institutionalized population of the United States. The survey is comprised of two overlapping panels that are each included in the survey for two years. Newborns and infants under 2 years old on December 31 of a survey year were included in the study. Additionally, adult men and women living in the same household between 18-45 years old were considered fathers and mothers of infants. Histamine2-receptor antagonists (H2RA) and proton pump inhibitors (PPI) were identified for infants. H2RA, PPIs, and anti-depressant (selective serotonin re-uptake inhibitors and serotonin-norepinephrine re-uptake inhibitors) were identified for parents. Sex, age (by month for child), race/ethnicity (White non-Hispanic, Black non-Hispanic, Hispanic, or other), region (Northeast, Midwest, South, and West), and poverty category were identified. Chi-squared and adjusted Wald tests were used to determine statistical significance with acid-suppressing medication use between 2010-2018. A multivariable logistic regression predicted any acid suppressing medication use with month of age and month of age2 as independent variables of use for the population between 2009-2018. The study was considered exempt by the OhioHealth Institutional Review Board. Survey weighting was included in all analyses. Results: The study included a total of 16,604 infants between 2002-2018. The rate of any acid suppressing medication increased between 2002-2004. H2RA use was more common than PPI use, especially after 2012. H2RA use appeared to increase after 2015. PPI use initially increased, but then remained relatively stable before appearing to decrease in 2015-2018. Ranitidine clearly had the most users of the H2RA, which was maintained throughout the study period. Lansoprazole had the most users among users of PPIs, but this decreased notably after 2010. Of the infants with a reported PPI, 34.0% (95%CI:23.6-46.3) also had an H2RA reported during the year. In total, 40.8% (95%CI:35.0-46.8) had only 1 reported fill of an acid suppresser. There were 8,075 infants under 2 years of age between 2010-2018. The rate of use was highest in infants between 4-11 months of age at the end of the survey year at 8.6% (95%CI:6.9-10.6). Acid-suppressing medication use was more common among infants in higher income families, White (non-Hispanic) race/ethnicity, infants with private health insurance, parents who reported acid-suppressing medication, maternal anti-depressant use, and certain regions of the country. Discussion: Between 6.9-10.6% of infants used an acid suppressing medication before their second birthday between 2010-2018, of which around 60% had multiple medication fills. It appears that there has been a small increases in use over the last few decades, of which ranitidine appears to be the medication driving the increase. Numerous socio-economic and demographic characteristics were associated with acid suppressing medication use. This study has numerous limitations, including potential under-reporting of acid-suppressing medications, an imperfect identification of parent, unreliable diagnoses (not included in the analysis), and a relatively small sample size. We opted not to run a multivariable logistic regression of socio-economic and demographic characteristics given concern for table 2 fallacy and relatively small sample size. Acid suppressing medication use was common in infants before their second birthday. Additional research should be conducted on efficacy and safety of these medications given the level of use and very low quality of available evidence.

## Dear Editor

I attempted to determine rates of use and associations with use of acid-suppressing medications in infants under 2 years old given these are not well studied.^1^

## Methods

The 2002-2018 Medical Expenditure Panel Survey (MEPS) was used for the analysis.^2^ The survey is sponsored by the Agency of Healthcare Research and Quality and is conducted on households to be nationally representative of the non-institutionalized population of the United States. The survey is comprised of two overlapping panels that are each included in the survey for two years.

Newborns and infants under 2 years old on December 31 of a survey year were included in the study. Additionally, adult men and women living in the same household between 18-45 years old were considered fathers and mothers of infants.

Histamine_2_-receptor antagonists (H_2_RA) and proton pump inhibitors (PPI) were identified for infants. H_2_RA, PPIs, and anti-depressant (selective serotonin re-uptake inhibitors and serotonin-norepinephrine re-uptake inhibitors) were identified for parents. Sex, age (by month for child), race/ethnicity (White non-Hispanic, Black non-Hispanic, Hispanic, or other), region (Northeast, Midwest, South, and West), and poverty category were identified.

Chi-squared and adjusted Wald tests were used to determine statistical significance with acid-suppressing medication use between 2010-2018. A multivariable logistic regression predicted any acid suppressing medication use with month of age and month of age^2^ as independent variables of use for the population between 2009-2018.

The study was considered exempt by the OhioHealth Institutional Review Board. Survey weighting was included in all analyses.

## Results

The study included a total of 16,604 infants between 2002-2018. The rate of any acid suppressing medication increased between 2002-2004. H_2_RA use was more common than PPI use, especially after 2012. H_2_RA use appeared to increase after 2015. PPI use initially increased, but then remained relatively stable before appearing to decrease in 2015-2018. (Figure 1, Left) Ranitidine clearly had the most users of the H_2_RA, which was maintained throughout the study period. Lansoprazole had the most users among users of PPIs, but this decreased notably after 2010. (Figure 1, Right) Of the infants with a reported PPI, 34.0% (95%CI:23.6-46.3) also had an H_2_RA reported during the year. In total, 40.8% (95%CI:35.0-46.8) had only 1 reported fill of an acid suppresser.

**Figure 1:**
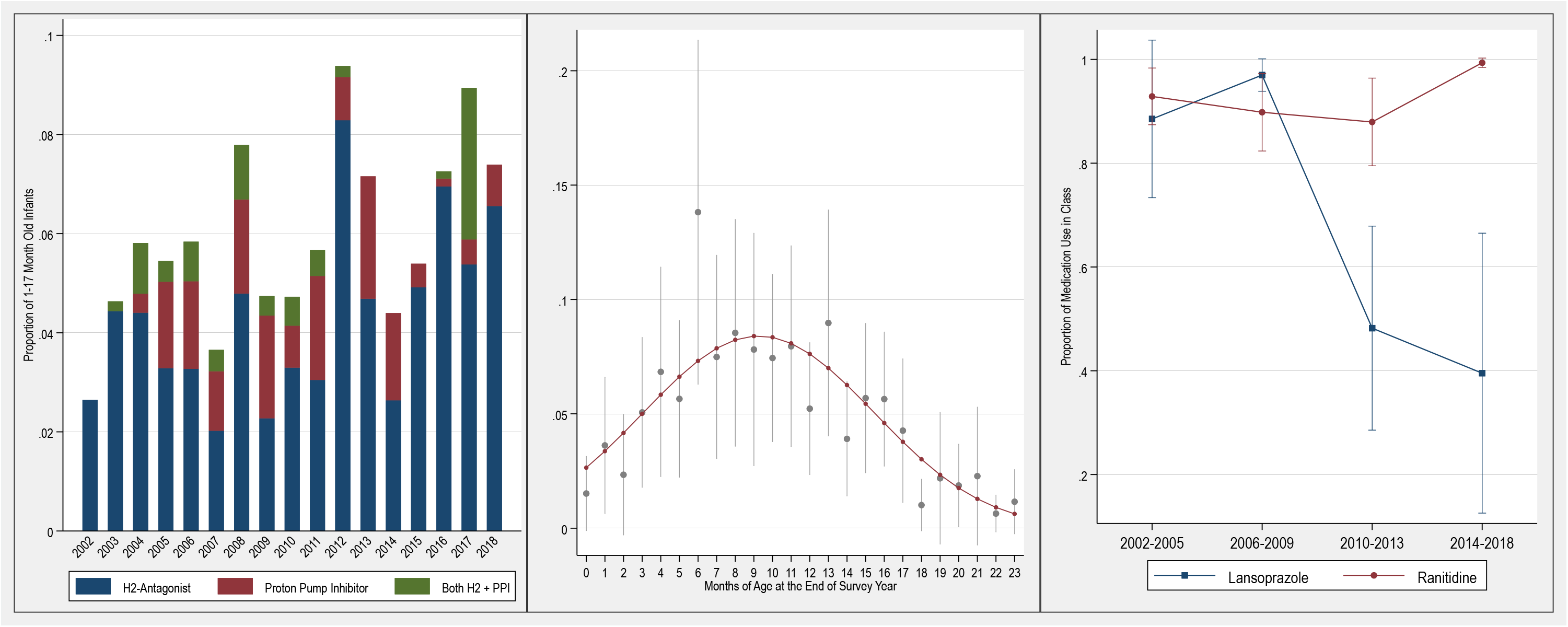
Left: Proportion of Infants who had a filled prescription for H_2_-antagonist, Proton Pump Inhibitor, and both. The figure includes only infants who were between 1 and 17 months old at the end of the survey year. Center: Proportion of Infants who had a filled prescription for an acid-suppressing agent between 2009-2018 by month age at the end of survey year. Fitted line is derived from a multivariable logistic regression with independent variables of months of age and months of age^2^. Right: The proportion of H_2_-antagonists users who reported a fill of ranitidine and the proportion of proton pump inhibitor users who reported a fill of lansoprazole.

There were 8,075 infants under 2 years of age between 2010-2018. The rate of use was highest in infants between 4-11 months of age at the end of the survey year at 8.6% (95%CI:6.9-10.6) (Figure 1, Center). Acid-suppressing medication use was more common among infants in higher income families, White (non-Hispanic) race/ethnicity, infants with private health insurance, parents who reported acid-suppressing medication, maternal anti-depressant use, and certain regions of the country. (Table 1)

**Table 1:** Bivariate Analysis of Acid-Suppressing Medication Use, 2010-2018. P-values represent differences with Adjusted Wald Test for continuous variables and chi-squared test for categorical variables.

|  | **No H2/PPI** | **H2 or PPI** | **P-Value** |
| --- | --- | --- | --- |
| **Any H2 or PPI** | 7802 | 273 |  |
| **Proportion of population** | 94.8 (0.5) | 5.2 (0.5) |  |
| **H2-Blocker (n)** | 95.6 (0.5) | 4.4 (0.5) |  |
| **Proton Pump Inhibitor (n)** | 98.8 (0.2) | 1.2 (0.2) |  |
| **Age (in Months, SE)** | 11.8 (0.09) | 9.8 (0.40) | <0.001 |
| **Sex, % Female (SE)** | 48.9 (0.1) | 50.1 (4.7) | 0.8 |
| **Race/Ethnicity, % (SE)** |  |  | <0.001 |
| White (Non-Hispanic) | 49.9 (1.5) | 72.7 (3.6) |  |
| Black (Non-Hispanic) | 13.0 (0.8) | 7.1 (1.5) |  |
| Hispanic | 26.5 (1.3) | 13.2 (2.9) |  |
| Other | 10.7 (0.7) | 7.0 (1.9) |  |
| **Poverty Category, % (SE)\*\*** |  |  | 0.002 |
| Poor/Near Poor | 31.5 (1.1) | 19.7 (2.7) |  |
| Low/Middle Income | 42.5 (1.0) | 40.6 (5.0) |  |
| High Income | 26.1 (1.1) | 39.7 (4.8) |  |
| **Insurance, % (SE)** |  |  | 0.009 |
| Any Private | 52.2 (1.2) | 64.1 (4.0) |  |
| Public | 42.7 (1.3) | 30.9 (4.0) |  |
| Uninsured | 5.2 (0.4) | 5.0 (1.5) |  |
| **Region, % (SE)** |  |  | 0.006 |
| Northeast | 15.4 (1.0) | 25.6 (4.5) |  |
| Midwest | 21.5 (1.1) | 25.4 (4.1) |  |
| South | 38.0 (1.4) | 32.7 (4.7) |  |
| West | 25.1 (1.2) | 16.3 (3.5) |  |
| **Maternal Acid Suppression** |  |  | 0.001 |
| None | 95.6 (0.4) | 85.8 (4.1) |  |
| Acid Suppression | 3.2 (0.3) | 12.0 (3.9) |  |
| No Mother Identified | 1.2 (0.2) | 2.2 (1.6) |  |
| **Paternal Acid Suppression** |  |  | 0.01 |
| None | 77.0 (0.8) | 80.3 (3.7) |  |
| Acid Suppression | 2.1 (0.3) | 5.8 (2.2) |  |
| No Father Identified | 20.9 (0.8) | 13.9 (3.3) |  |
| **Maternal Anti-Depressant Use** |  |  | 0.02 |
| None | 91.2 (0.5) | 83.0 (3.6) |  |
| Anti-Depressant Use | 7.6 (0.5) | 14.8 (3.3) |  |
| No Mother Identified | 1.2 (0.2) | 2.2 (1.6) |  |
| **Paternal Anti-Depressant Use** |  |  | 0.1 |
| None | 76.4 (0.8) | 84.2 (3.4) |  |
| Anti-Depressant Use | 2.7 (0.4) | 1.9 (0.8) |  |
| No Father Identified | 20.9 (0.8) | 13.9 (3.4) |  |

\*\*Excludes individuals from 2018 as data was not available at the time of analysis.

## Discussion

Between 6.9-10.6% of infants used an acid suppressing medication before their second birthday between 2010-2018, of which around 60% had multiple medication fills. It appears that there has been a small increases in use over the last few decades, of which ranitidine appears to be the medication driving the increase. Numerous socio-economic and demographic characteristics were associated with acid suppressing medication use.

This study has numerous limitations, including potential under-reporting of acid-suppressing medications, an imperfect identification of parent, unreliable diagnoses (not included in the analysis), and a relatively small sample size. We opted not to run a multivariable logistic regression of socio-economic and demographic characteristics given concern for table 2 fallacy and relatively small sample size.^3^

Acid suppressing medication use was common in infants before their second birthday. Additional research should be conducted on efficacy and safety of these medications given the level of use and very low quality of available evidence.^4-5^

## Data Availability

Data is publicly available through the MEPS website.

## Acknowledgments

I would like to thank Raj Mehta, MD for his comments on the the manuscript.

## References

1. Illueca M, Alemayehu B, Shoetan N, Yang H. Proton pump inhibitor prescribing patterns in newborns and infants. J Pediatr Pharmacol Ther. 2014;19(4):283–287. doi:10.5863/1551-6776-19.4.283

2. Medical Expenditure Panel Survey: 2017 Full Year Consolidated Data File. https://meps.ahrq.gov/data_stats/downloaddata/pufs/h201/h201doc.pdf. Accessed August 4, 2020.

3. Westreich D, Greenland S. The table 2 fallacy: presenting and interpreting confounder and modifier coefficients. Am J Epidemiol. 2013;177(4):292–298. doi:10.1093/aje/kws412

4. Rosen R, Vandenplas Y, Singendonk M, et al. Pediatric Gastroesophageal Reflux Clinical Practice Guidelines: Joint Recommendations of the North American Society for Pediatric Gastroenterology, Hepatology, and Nutrition and the European Society for Pediatric Gastroenterology, Hepatology, and Nutrition. J Pediatr Gastroenterol Nutr. 2018;66(3):516–554. doi:10.1097/MPG.0000000000001889

5. Hassall E. Over-prescription of acid-suppressing medications in infants: how it came about, why it’s wrong, and what to do about it. J Pediatr. 2012;160(2):193-198. doi:10.1016/j.jpeds.2011.08.067

